# Factors associated with non-use of ART among men living with HIV in South Africa: findings from a population-based household survey

**DOI:** 10.1101/2023.05.04.23289507

**Authors:** Nuha Naqvi, Xan Swart, Jeremiah Chikovore, Kassahun Ayalew, Sizulu Moyo, Diane Morof, David Makapela, Jason Bedford

## Abstract

**Introduction:** Although South Africa adopted the World Health Organization’s Test and Treat strategy for HIV epidemic control in 2016, antiretroviral therapy (ART) treatment initiation and retention remain below target. In 2017, an estimated 56.3% of men living with HIV were on ART. We aimed to investigate factors associated with non-use of ART among men in South Africa.

**Methods:** Utilizing data from the fifth South African National HIV Prevalence, Incidence, Behavior and Communication (SABSSM V) cross-sectional survey conducted in 2017, a subset of data from HIV-positive men was stratified based on presence/absence of antiretroviral drugs (ARVs) detected in dried blood spot samples. Data were weighted to be representative of the national population and analyzed using multivariable logistic regression to assess predictors of non-use of ART; p<0.05 was considered significant.

**Results:** A total of 6,920 men aged ≥15 years old were enrolled in the study, of whom 953 (13.8%) tested HIV-positive. Among those HIV-positive, 810 (85%) had a known ARV test result: 470 (58%) had ARVs detected, and 340 (42%) did not have ARVs detected. Non-use of ART in men was associated with high-risk alcohol use (adjusted odds ratio (AOR)=3.68, 95% confidence interval (CI): 1.03-13.23), being a widower compared to being unmarried (AOR=6.99, 95%CI: 1.46-33.42), and having drug-resistant HIV (AOR=28.12, 95%CI: 13.89-56.94). Per year increase in age (AOR=0.67, 95%CI: 0.47-0.96), residence in rural tribal localities compared to urban localities (AOR=0.38, 95%CI: 0.18-0.78), or having a co-morbidity such as tuberculosis or diabetes (AOR=0.06, 95%CI: 0.03-0.14) were positively associated with ART use.

**Conclusions:** Non-use of ART was strongly associated with HIV drug resistance. Young men who are living with HIV, those with high-risk alcohol use, and widowers, should be a priority for HIV programming and linkage to care. Identifying interventions that are effective at linking these men to ART will help reduce the burden of HIV in South Africa.

## Introduction

Despite notable gains made in recent years to curb new infections, South Africa continues to battle the world’s largest HIV epidemic. Although South Africa adopted the World Health Organization’s Test-and-Treat strategy for HIV epidemic control in 2016 to make antiretroviral therapy (ART) more accessible and available at no cost in the public sector, treatment initiation and retention remain below target. Poor retention and non-ART use increases the risk of suboptimal viral suppression, and consequently the likelihood of further transmission of HIV, drug resistance, treatment failure, associated morbidity and mortality.

The fifth South Africa National HIV Prevalence, Incidence, Behavior and Communication (SABSSM V) survey, conducted in 2017 found that 56.3% and 65.5% of men and women living with HIV, respectively, were on ART [1]. The revised Joint United Nations Programme on HIV/AIDS (UNAIDS) Fast-Track 95– 95–95 targets aim to ensure that 95% of people living with HIV (PLHIV) are tested and diagnosed, 95% of people diagnosed with HIV are receiving treatment, and 95% of people on treatment have a suppressed viral load. By 2019, estimates indicated that in South Africa, 90% of people were aware of their HIV status while only 68% of those diagnosed with HIV were on ART [2]. In 2021, ART coverage was estimated at only 61.9% among males and 72.3% among females aged 15 years and older [3].

Across sub-Saharan Africa, men test for HIV at lower rates, have higher rates of attrition from treatment programs, higher rates of virologic failure on ART, and higher mortality on ART [4]. Men are generally less likely to engage and remain in care compared to women [5-7], and are more likely to experience a deterioration in health after an HIV diagnosis [8]. Traditional requirements of men’s employment and livelihoods tend to contribute to sub-optimal testing and engagement in treatment. A feeling of shame for appearing “weak” is a well-documented deterrent for men when accessing treatment and healthcare services [9]. Many men also fear being held responsible for transmitting HIV to their female sexual partners, experiencing discomfort or embarrassment at being seen in health spaces that are considered feminised spaces, and fear that engaging in care threatens their social standing and ability to socialise or engage in work [10]. Entrenched gender norms promulgating the view that care-seeking and healthcare spaces are primarily the woman’s domain, contribute to an avoidance of HIV treatment among men. In addition to stigma related to seeking healthcare, higher rates of alcohol and substance use among men are also determinants of poor adherence to ART [11-12].

It is increasingly clear that strategies to improve access to care and health outcomes among men living with HIV are essential to reach the UNAIDS 95-95-95 targets by 2030. The objective of this analysis is to investigate factors associated with non-use of ART among men in South Africa. We describe uptake of ART uptake among men who tested HIV-positive in the 2017 SABSSM V survey, and explore barriers associated with effective linkage to HIV care and treatment for men.

## Methods

The data used in this paper were obtained from SABSSM V, a cross-sectional, population-based household HIV survey conducted in 2017 in South Africa. The multistage stratified random cluster sampling approach of the survey is fully described elsewhere [1]. In summary, 1,000 small area layers (SALs) were selected and stratified by province, locality type, and race groups to be nationally representative. A maximum of 15 households/visiting points were randomly selected from each SAL. All household members who slept in the selected household the previous night were eligible to participate. After obtaining consent, household level data were collected through a questionnaire administered to the head of the household. Thereafter, individual consent was obtained and an individual questionnaire that collected sociodemographic information, knowledge and attitudes about HIV, and sexual behavior data was administered. Adults aged 18 years or older signed informed consent forms, while parental/legal guardian consent and individual assent were obtained for those aged 15-17 years old. The survey questionnaires were captured electronically on a Mercer A105 tablet utilizing Census and Survey Processing System (CSPro) software.

Dried blood spot (DBS) specimens were collected by finger prick to test for HIV status, viral load (VL), HIV recency using the limiting-antigen (LAg) avidity enzyme immunoassay (EIA), HIV drug resistance (HIVDR), and detection of antiretroviral drugs (ARVs) from consenting participants. HIV antibody status was determined using an algorithm with two different EIAs and a nucleic acid amplification test (NAAT) for validating all positive results. VL testing used the Abbott platform (Abbott m2000 HIV Real-Time System, Abbott Molecular Inc., Des Plaines, IL, USA). Samples with VL >1,000 copies/mL were classified as virally unsuppressed and were submitted for HIVDR testing. HIVDR testing was conducted by Next Generation Sequencing (NGS). Detection of ARVs in DBS samples was done by high-performance liquid chromatography (HPCL) coupled with Tandem Mass Spectrometry. The testing panel focused on drugs used in the public sector programme, and was set to detect Nevirapine, Efavirenz, Lopinavir, Atazanavir and Darunavir. The detection limit was set at 0.02 μg/mL for each drug, with a signal-to-noise ratio of at least 5:1.

The Human Sciences Research Council Research Ethics Committee provided ethical approval for the survey (REC 4/18/11/15). This project was reviewed in accordance with Centers for Disease Control and Prevention (CDC) human research protection procedures and was determined to be research, but CDC investigators did not interact with study participants or have access to identifiable data or specimens for research purposes. Personal identifiable information was not collected in the survey and confidentiality was maintained during data collection, storage, analysis, and reporting.

For this analysis, a sub-sample comprised of HIV-positive male participants ≥15 years of age was extracted. Female participants, male participants <15 years of age, and those who tested HIV-negative at the time of the survey were excluded from this analysis. The outcome of interest was ARV use – those in whom *ARVs were detected* (one or more of the five drugs in the testing panel) and those in whom *ARVs were undetected*, representing males who were currently taking ARVs and males who were not (the latter includes both those that were ART non-adherent and those that had never initiated ART) at the time of sample collection. Figure 1 shows a flow diagram of study participants included in this analysis.

**Figure 1:**
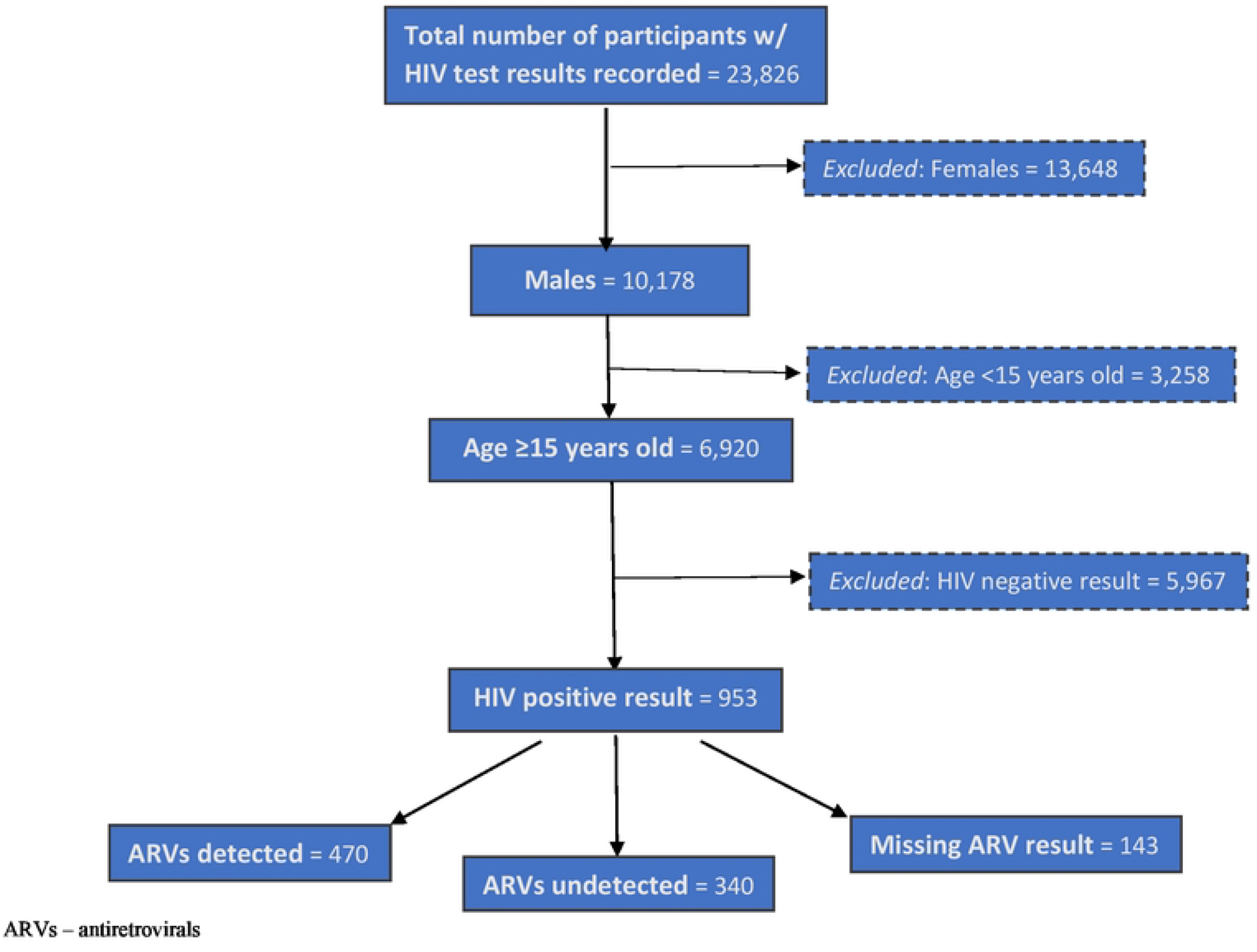
Study Sample Flow Diagram, SABBSM V survey, 2017.

### Variables

In this analysis, ART use is defined based on the detection of any ARVs in blood specimens. Although ART adherence was verbally volunteered by participants, these data were not used in this analysis due to possible social desirability bias (being non-adherent is seen as a less desirable trait) and the authors opted for a more robust observation of ART use.

Probing factors associated with ART non-use in South African HIV-positive males followed a multi-faceted approach. Behavioral and demographic variables with known relationships to ART uptake and other variables that were significant in the univariate analysis were then built into a logistic regression model. Multi-level nominal variables with small samples were merged into variable categories for concise analysis. Employment status, for example, originally consisted of five categories including 1) unemployed, 2) sick/disabled and unable to work, 3) student/pupil, 4) employed/self-employed, and 5) other. This was reduced to *employed* or *unemployed* groupings, which aided in generating relatively stable estimates in the regression analysis. This reduction approach was used for level of education, main source of healthcare, number of comorbidities, history of drug use, and number of life-time sexual partners.

Participants who scored 16 or more on the Alcohol Use Disorders Identification Test (AUDIT) [13] tool were labelled as high risk for alcohol abuse/addiction in this study.

### Analysis

All statistical analyses were conducted using STATA software version 14.0 (Stata Corporation, College Station, TX, United States) and RStudio 4.1.2 [14] using the survey package 4.0 [15], with data weighted to be representative of the national population by sex, age, and race for 2017 as provided by the SABSSM V survey. Exploratory analysis of factors assessed the variables’ respective distributions followed by appropriate hypothesis tests (student’s t-test, chi-squared, Wilcoxon Rank-Sum tests) to test for statistically significant differences between the detected and undetected ARV groups. Missing values were excluded and p < 0.05 was considered significant.

Univariate logistic regression was used to analyze the strength of associations between individual predictor variables and outcome variables, reported as odds ratios (ORs). A multivariable logistic regression model including all relevant behavioral and sociodemographic variables was constructed to measure the net effect of predictor variables on the outcome of ART non-use. Continuous independent variables were standardized and centered at their respective means. Assessment of model fit parameters (Akaike Information Criterion (AIC), pseudo-R^2^) and level of missingness in certain factors guided the construction of a final reduced multivariable logistic regression model, the results of which, are reported as adjusted odds ratios (AORs) and 95% confidence intervals (CIs).

## Results

A total of 6,920 men aged ≥15 years were enrolled in the study, of whom 953 (13.8%) had a laboratory confirmed HIV-positive result. Among those HIV-positive, 810 (85.0%) had a known ARV test result: 470 (58%) had ARVs detected, and 340 (42%) did not have ARVs detected. Table 1 shows the unweighted frequencies and weighted proportions of sociodemographic, behavioral, and clinical characteristics of participants by ARV status. The mean age of the total study population was 38 years old, lower among the ARV-undetected group (mean age of 34 years old) than the ARV-detected group (mean age of 41 years old). Most participants identified as Black African (93.9%) and resided in urban areas (67.4%). More than one quarter (29%) graduated from high school/finished matric and approximately half were unemployed (50.6%). Most participants reported never being married (69.1%); a greater proportion of those in the ARV-undetected group (74.7%) reported never being married as compared to the ARV-detected group (64.5%).

**Table 1.** Sociodemographic and behavioral characteristics of HIV-positive men aged ≥15 years by ARV status, 2017 SABSSM V survey.

|  | **Overall**<br>**(N=810)** | **ARVs detected**<br>**(n=470)** | **ARVs**<br>**undetected**<br>**(n=340)** |
| --- | --- | --- | --- |
|  | *Mean (SD)* | *Mean (SD)* | *Mean (SD)* |
| **Age (years)** | 38 (11.05) | 41 (11.47) | 34 (9.25) |
|  | *N (weighted %)* | *n (weighted %)* | *n (weighted %)* |
| **Locality type** |  |  |  |
| Urban | 441 (67.4) | 243 (63.3) | 198 (72.4) |
| Tribal Rural | 269 (26.6) | 181 (32.1) | 88 (19.9) |
| Farm Rural | 100 (6.0) | 46 (4.6) | 54 (7.7) |
| **Race** |  |  |  |
| Black African | 654 (93.9) | 382 (94.2) | 272 (93.5) |
| Coloured | 55 (4.4) | 26 (3.3) | 29 (5.7) |
| Indian/White | 30 (1.7) | 18 (2.5) | 12 (0.8) |
| **Marital Status** |  |  |  |
| Married | 176 (24.5) | 113 (28.0) | 63 (20.3) |
| Never Married | 542 (69.1) | 301 (64.5) | 241 (74.7) |
| Divorced / separated | 25 (3.3) | 19 (4.4) | 6 (1.9) |
| Widower | 28 (3.1) | 16 (3.2) | 12 (3.1) |
| **Employment Status = Unemployed** | 426 (50.6) | 277 (57.2) | 149 (42.6) |
| **Level of Education Achieved** |  |  |  |
| No Education | 65 (6.8) | 43 (8.1) | 22 (5.3) |
| Completed Primary School | 182 (19.0) | 118 (21.8) | 64 (15.7) |
| Completed Grade 8-11 | 330 (44.4) | 185 (44.1) | 145 (44.8) |
| Completed Matric or Higher Education | 193 (29.7) | 102 (26.0) | 91 (34.2) |
| **Perception of own health** |  |  |  |
| Excellent | 166 (21.6) | 90 (19.2) | 76 (24.6) |
| Good | 400 (55.5) | 218 (51.1) | 182 (61.0) |
| Fair | 159 (17.6) | 108 (23.1) | 51 (10.8) |
| Poor | 43 (5.3) | 31 (6.6) | 12 (3.6) |
| **Time since last visit to health facility** |  |  |  |
| Within the past six months | 415 (55.3) | 292 (66.1) | 123 (42.0) |
| More than six months but not more than a year ago | 105 (12.1) | 55 (10.4) | 50 (14.2) |
| More than one year ago | 172 (22.8) | 69 (15.6) | 103 (31.6) |
| Never | 75 (9.8) | 30 (7.9) | 45 (12.3) |
| **Main source of healthcare** |  |  |  |
| Government | 677 (85.3) | 406 (86.7) | 271 (83.6) |
| Private | 74 (11.4) | 37 (11.4) | 37 (11.4) |
| Other | 15 (3.3) | 3 (1.9) | 12 (5.1) |
| **Previous TB diagnosis = No** | 620 (82.2) | 325 (73.0) | 295 (93.5) |
| Yes | 190 (17.8) | 145 (17.0) | 45 (6.5) |
| **Comorbidities** |  |  |  |
| No comorbidities | 495 (64.0) | 208 (45.9) | 287 (86.4) |
| One Comorbidity | 243 (28.2) | 201 (41.6) | 42 (11.5) |
| Two or More Comorbidities | 72 (7.8) | 61 (12.4) | 11 (2.0) |
| **Presence of a disability: Yes** | 43 (4.5) | 32 (6.8) | 11 (1.6) |
| **History of drug use = Yes** | 146 (22.5) | 78 (24.3) | 68 (20.1) |
| **High probability of alcohol abuse = Yes** | 46 (6.8) | 23 (5.5) | 23 (8.4) |
| **High risk alcohol use score (mean (SD))** | 3.44 (6.40) | 2.88 (5.81) | 4.22 (7.01) |
| **HIV drug resistance = Yes** | 332 (50.5) | 83 (22.6) | 249 (83.6) |
| **Number of lifetime sexual partners** |  |  |  |
| Zero/one partner | 81 (13.2) | 44 (12.9) | 37 (13.7) |
| Two-Three Partner | 116 (15.5) | 66 (14.6) | 50 (16.7) |
| More than three partner | 389 (71.3) | 235 (72.5) | 154 (69.6) |
| **Previous sexually transmitted infection** | 49 (9.6) | 27 (8.2) | 22 (11.2) |
| **Demonstrated understanding of HIV prevention = Yes** | 467 (62.3) | 279 (61.6) | 188 (63.1) |
| **Demonstrated understanding of HIV treatment = Yes** | 571 (79.3) | 359 (87.3) | 212 (69.3) |
| **Stigma towards HIV positive individuals = Yes** | 334 (40.2) | 178 (38.9) | 156 (41.7) |
| **Has a mental illness or exhibits mental distress = Present** | 329 (43.1) | 192 (43.3) | 137 (42.8) |

Table 1 includes the unweighted frequencies (N/n) and weighted proportions (%) of sociodemographic, behavioral, and clinical characteristics of participants by ARV status.
Abbreviations: ART – Antiretroviral therapy; SABSSM – South Africa National HIV Prevalence, Incidence, Behavior and Communication survey; SD – standard deviation; TB – tuberculosis.

Table 2 displays the univariate and multivariable logistic regression models of factors associated with non-use of ART. In observing the univariate model, being employed was found to be a predictor of ART use compared to those who are unemployed (odds ratio (OR)=0.56, 95%CI: 0.37-0.83, p<0.001). A demonstrated understanding of HIV treatment was found to be associated with a decreased likelihood of ART non-use (OR=0.33, 95%CI: 0.19-0.56, p<0.001). Presence of a disability decreased the likelihood of ART non-use among men compared to the absence of disability (OR=0.22, 95%CI: 0.07-0.68, p=0.010). Having no history of a tuberculosis (TB) diagnosis significantly increased the odds of not using ART (OR=5.30, 95%CI: 2.55-11.01, p<0.001). However, these variables were not determined to be significant in the multivariable models.

**Table 2:** Factors associated with ART non-use among men aged ≥15 years, 2017 SABSSM V survey.

|  | **Univariate Model** | **Saturated Multivariable Model** | **Reduced Multivariable Model** |
| --- | --- | --- | --- |
|  | *OR (95%CI) p-value* | *aOR (95%CI) p-value* | *aOR (95%CI) p-value* |
| **Age (years)** | **0.51 (0.41-0.62) <0.001** | **0.41 (0.24-0.69) <0.001** | **0.67 (0.47-0.96) 0.030** |
| **Locality type of residence** |  |  |  |
| Urban | *1 (reference)* | *1 (reference)* | *1 (reference)* |
| Tribal Rural | **0.54 (0.35-0.84) 0.010** | **0.26 (0.09-0.72) 0.010** | **0.38 (0.18 0.78) 0.010** |
| Farm Rural | 1.46 (0.83-2.59) 0.190 | 0.47 (0.07-3.16) 0.440 | 0.83 (0.30-2.30) 0.720 |
| **Race** |  |  |  |
| Black African | *1 (reference)* | *1 (reference)* | *1 (reference)* |
| Coloured | 1.74 (0.87-3.48) 0.120 | 0.45 (0.04-4.99) 0.510 | 1.41 (0.41-4.67) 0.610 |
| Indian/White | **0.33 (0.12-0.91) 0.030** | **0.07 (0.005-0.978) 0.049** | **0.21 (0.06-0.84) 0.027** |
| **Marital Status** |  |  |  |
| Never Married | *1 (reference)* | *1 (reference)* | *1 (reference)* |
| Married | 0.63 (0.38-1.02) 0.060 | 2.94 (0.75-11.48) 0.120 | 2.24 (0.873-5.73)0.090 |
| Divorced / separated | 0.38 (0.10-1.46) 0.160 | 3.08 (0.58-16.40) 0.190 | 0.56 (0.14-2.28) 0.423 |
| Widower | 0.84 (0.26-2.70) 0.780 | 11.11 (0.29-424.52) 0.200 | **6.99 (1.46-33.42) 0.020** |
| **Employment Status = Employed** | **0.56 (0.37-0.83) <0.001** | 0.49 (0.18-1.32) 0.160 | 0.78 (0.38-1.62) 0.510 |
| **Number of comorbidities** |  |  |  |
| No comorbidities | *1 (reference)* | *1 (reference)* | *1 (reference)* |
| One Comorbidity | **0.15 (0.09-0.25) <0.001** | **0.10 (0.03-0.29) <0.001** | **0.06 (0.03-0.14) <0.001** |
| Two or More Comorbidities | **0.09 (0.03-0.23) <0.001** | **0.13 (0.02-0.73) 0.020** | **0.04 (0.01-0.13) <0.001** |
| **High probability of Alcohol Abuse** |  |  |  |
| High probability of Alcohol abuse | 1.60 (0.71-1.13) 0.260 | **4.43 (1.27 -15.47) 0.020** | **3.68 (1.03-13.23) 0.046** |
| **HIV drug resistance** |  |  |  |
| Present | **17.45 (10.02-30.39) <0.001** | **30.40 (10.01-92.32) <0.001** | **28.12 (13.89-56.94) <0.001** |
| **Presence of a Disability:** |  |  |  |
| Disability present | **0.22 (0.07-0.68) 0.010** | 0.11 (0.01-1.08) 0.060 | 0.16 (0.02-1.11) 0.064 |
| **Level of Education Achieved** |  |  |  |
| No Education | *1 (reference)* | *1 (reference)* |  |
| Completed Primary School | 1.10 (0.46-2.63) 0.830 | 0.71 (0.14-3.70) 0.690 |  |
| Completed Grade 8-11 | 1.55 (0.68-3.52) 0.290 | 0.59 (0.14-2.47) 0.470 |  |
| Completed Matric or Higher Education | 2.01 (0.85-4.73) 0.110 | 0.81 (0.15-4.44) 0.810 |  |
| **Perception of participant's own health** |  |  |  |
| Excellent | *1 (reference)* | *1 (reference)* |  |
| Good | 0.93 (0.56-1.55) 0.790 | 2.81 (0.76-10.43) 0.120 |  |
| Fair | **0.37 (0.19-0.70) <0.001** | 2.61 (0.56-12.26) 0.220 |  |
| Poor | 0.42 (0.14-1.31) 0.140 | 0.72 (0.04-13.02) 0.820 |  |
| **Participant's time since previous health visit** |  |  |  |
| Within the past six months | *1 (reference)* | *1 (reference)* |  |
| More than six months but not more than a year ago | **2.14 (1.18-3.88) 0.010** | 1.52 (0.43-5.36) 0.510 |  |
| More than one year ago | **3.18 (1.87-5.39) <0.001** | 0.47 (0.10-2.26) 0.350 |  |
| Never | **2.45 (1.25-4.83) 0.010** | 0.84 (0.17-4.28) 0.840 |  |
| **Participant's main source of health care** |  |  |  |
| Government | *1 (reference)* | *1 (reference)* |  |
| Private | 1.03 (0.53-2.00) 0.920 | 1.22 (0.08-19.21) 0.890 |  |
| Other | 2.84 (0.64-12.58) 0.170 | 3.19 (0.06-179.46) 0.570 |  |
| **Previous TB diagnosis** |  |  |  |
| No previous diagnosis | **5.30 (2.55-11.01) <0.001** | 2.82 (0.53-14.88) 0.220 |  |
| **Participant has a history of drug use** |  |  |  |
| Participant admits to using drugs | 0.78 (0.46-1.32) 0.360 | 0.93 (0.32-2.72) 0.900 |  |
| **Previous sexually transmitted infection** |  |  |  |
| Has had a previous STI | 1.41 (0.63-3.16) 0.400 | 1.25 (0.21-7.36) 0.810 |  |
| **Demonstrated understanding of HIV prevention** |  |  |  |
| Has demonstrated a basic understanding | 1.07 (0.70-1.63) 0.750 | 0.67 (0.24-1.87) 0.450 |  |
| **Demonstrated understanding of HIV treatment** |  |  |  |
| Has demonstrated a basic understanding | **0.33 (0.19-0.56) <0.001** | 0.47 (0.08-2.78) 0.400 |  |
| **Exhibits stigma towards HIV positive individuals** |  |  |  |
| Exhibited Stigma towards HIV positive people | 1.12 (0.75-1.68) 0.570 | 1.27 (0.52-3.10) 0.600 |  |

Table 2 includes the univariate and multivariable logistic regression (saturated and reduced) models of factors associated with non-use of ART. Odds ratios, confidence intervals, and p-values are listed for the univariate model. Adjusted odds ratios, confidence intervals, and p-values are listed for the multivariable models.
Abbreviations: A/OR – adjusted/odds ratio; ART – Antiretroviral therapy; SABSSM – South Africa National HIV Prevalence, Incidence, Behavior and Communication survey;
TB – tuberculosis.

Adjusting for age (and other known covariates) in the reduced multivariable model, non-use of ART in men was associated with high risk of alcohol abuse (adjusted odds ratio (AOR)=3.68, 95%CI: 1.03-13.23, p=0.046), being a widower compared to being unmarried (AOR=6.99, 95%CI: 1.46-33.42, p=0.020), and having drug-resistant HIV (AOR=28.12, 95%CI: 13.89-56.94, p<0.001). Being in the Indian/white race group decreased the likelihood of ART non-use (AOR=0.21, 95%CI: 0.06-0.84, p=0.027).

The reduced multivariable model showed older age to be a significant predictor of ART use, with an AOR of 0.67 per year increase over the age of 38 years (AOR=0.67, 95%CI: 0.47-0.96, p=0.030) for non-use of ART. Residence in rural tribal localities compared to urban localities (AOR=0.38, 95%CI: 0.18-0.78, p=0.010) and having a co-morbidity such as diabetes or cardiac disease (AOR=0.06, 95%CI: 0.03-0.14, p<0.001) were positively associated with ART use. Similarly, having two or more comorbidities reduced the adjusted odds of ART non-use by 0.04 (95% CI: 0.01-0.13, p-value <0.001) as compared to having no comorbidities.

Age, number of comorbidities, race, type of locality and HIV drug resistance remained statistically significant across the univariate, saturated, and reduced logistic regression models. High risk of alcohol abuse (AUDIT score of 16 or more) became significant in the reduced and saturated models but was not found in the univariate model. Being a widower gained statistical significance in the reduced model only.

The saturated model significantly reduced the cohort’s size due to missingness in some key independent predictors, such as lifetime sexual partners and demonstrated basic understanding of HIV treatment principles, that were being studied. This resulted in some variables being excluded in the reduced logistic regression model based on missingness. The reduced model, controlling for a number of predictors simultaneously, was selected based on AIC, Cragg-Uhler and McFadden pseudo-R^2^ model fit parameters. Previous history of a TB diagnosis, education level obtained, time since previous health visit did not improve the model fit criteria. The reduced model’s AIC was 413.1, McFadden and Cragg-Uhler’s pseudo-R^2^ was 0.51 and 0.68 respectively.

## Discussion

The UNAIDS 95–95–95 targets aim to ensure that 95% of people diagnosed with HIV are receiving sustained antiretroviral treatment. However, more than 42% of men living with HIV were not on ART at the time of data collection for this study. In this analysis ART use was higher among men above 38 years old, men in tribal rural areas compared to urban areas, and men with at least one previously known comorbidity. Non-use of ART in men was seen with high-risk alcohol use and was strongly associated with HIV drug resistance.

Men generally delay engaging with healthcare until their health has deteriorated severely [9]. The reasons for delay are largely related to how men internalize and deploy, within their lives and contexts, concepts of masculinity. Other reasons include lack of time to seek care (due to prioritizing earning and providing for families) or opting/feeling compelled to spend resources on basic requirements other than health [16]. However, this analysis found that men living with HIV with comorbidities are more likely to take up ART than those without known comorbidities. This may be because men with comorbidities are already familiar with healthcare facilities or have developed rapport with providers by seeking care for other conditions – which in turn impacts their behavior related to ART initiation and continuation. Co-morbidities can exacerbate or threaten to exacerbate experience of illness, and thus can serve to compel earlier engagement with healthcare. Further investigation and qualitative studies are needed to explore such hypotheses.

The positive association of ART use with increasing age aligns with findings regarding behavioral and biological indicators for HIV where younger age is generally associated with a higher viral load [17], which is a proxy for poor linkage to and uptake of care. The SABSSM V study also reported higher undiagnosed HIV but also comparatively lower HIV prevalence in younger age groups of men [1]. Modelling estimates for 2021 show HIV prevalence in men peaking in middle-aged (aged 35-44 years old), and an epidemiological shift of the burden to older men annually [3].

Barriers to accessing health services among young people are well-documented in the literature, and include prohibitive societal values [18-21], judgmental attitudes of health workers, limited access to relevant information, transport costs, and unsuitable facility operating hours [22-24]. Comparatively, older people are likely to have stable or wider social networks including larger and more elaborate family ties, facilitating financial or moral support to their engagement with healthcare [25]. Younger men are also more likely to engage in risky socialization activities that include or involve substance use. A qualitative study found a dominant pattern of decreasing binge and frequent drinking as men reached middle-age which was precipitated by family-building, reductions in drinking with work colleagues, and health concerns [25].

The present analysis also found a significant association between high-risk alcohol abuse and non-uptake of ART among other poor health outcomes, which is consistent with existing literature regarding alcohol and substances use and engagement with healthcare [9,17]. Alcohol and substance use are well documented as barriers to ART uptake [26], especially among younger men who tend to demonstrate ‘masculine bravado’, as part of assimilating to social groups or fitting in to society. This goes hand in hand with demonstrating invulnerability and repudiation of the stigmatized ‘unwell’ image [9,16,27].

The current analysis found that men residing in tribal informal areas were more likely to be on ART. A similar finding is reported from an analysis of the association between ART adherence and mental distress in South Africa, where PLHIV in rural/traditional localities were more likely to be adherent compared to those in urban areas [12]. Rural tribal areas had the highest response rate to the survey and demand for circumcision among adult men, but also the lowest in HIV testing, accurate HIV knowledge, and self-perceived ability to avoid HIV infection [1]. This makes the picture for rural settings complex and calls for a more in-depth analysis.

The demarcation ‘rural’ in South Africa is considered fluid and may vary across the country, thus its broad usage alongside ‘urban’ may mask finer patterns [28]. Provinces with large urban areas generally fare better in health outcomes including maternal mortality ratio, infant mortality rate, TB cure rates, and HIV burden [1] but also tend to have higher migration and mobility. Conversely, poorer health outcomes in rural areas are attributed to socio-economic indicators that include low education level, poor sanitation and unavailability of potable water, low household incomes, and food insecurity [27]. People in rural areas generally depend on public healthcare facilities, which typically are distant, and have limited staff, equipment, and packages of care [29]. Higher ART use in this analysis could be explained by the fact that informal tribal settings typically contain networks with more intimate connections and ties of community kinship, which may improve the rapport between patients and healthcare providers where more personal relationships have been established [30].

The finding that Indian and White populations were more likely to be on ART is consistent with HIV rates reported for these racial groups compared to Black Africans and Coloreds [1]. Race groups in South Africa tend to use different health sectors [31], and challenges with public healthcare services can be traceable to the segregationist policies of *apartheid* [32]. Since the end of *apartheid*, more doctors shifted to the private sector which limits access and affordability of care. Considering that various socio-economic advantages and vulnerabilities are conferred by race, these contextual and structural challenges affect ART utilization.

Aspects of social organization, including relative absence of anonymity, have significance where studies have illuminated concerns that providers routinely disclose health details of clients publicly in the community [33]. Not only do such actions restate the power imbalance between providers/healthcare systems and patients [34-35]; they also potentially drive HIV stigma. Studies from Southern Africa, for instance, report on patients not initiating HIV treatment due to the stigma of being witnessed using local clinics [36], or men being conscious of being labelled for using primary care facilities, which are held to be feminine spaces [37]. Substantial investment in the public HIV programs in South Africa could be instrumental in how the public healthcare system manages HIV care more efficiently. Increasing access to multi-month dispensing (MMD) of ART and using differentiated models of HIV care (DMOC) can minimize congestion in ART facilities and provide tailored ART care to support treatment continuity in men while reducing stigma and barriers to care [38].

Of great concern, this analysis found unacceptably high levels of drug resistance in men with ART non-use which may indicate ongoing transmission of drug resistant strains. As previously published, the SABSSM V survey found HIVDR mutations in 29.4% of males aged 15-49 years [39]. Resistance was detected in 27.4% of samples sequenced from virally unsuppressed respondents, and was most prevalent in samples from respondents with prior exposure to ART but no ARVs detected at the time of the survey [39]. Detecting this level of drug resistance is critical to ensuring that the ART regimens in use are optimal for the population. Dolutegravir-based regimens were not in use at the time of the SABSSM V survey, and these findings reinforce how critical the Tenofovir, Lamivudine, and Dolutegravir (TLD) transition continues to be. This underscores the importance of ART optimization and ensuring treatment adherence is strengthened among PLHIV. Men who have not initiated or have discontinued ART could be considered a priority population for initiation (or re-initiation) of optimized ART regimens to ensure that further resistance does not develop or spread, and a higher proportion of PLHIV are able to achieve viral suppression.

Given that Dolutegravir-based regimens (with a higher genetic barrier to resistance than non-nucleoside reverse transcriptase inhibitor (NNRTI)-based regimens) are now more widely used, findings from SABSSM VI and other population studies will be informative in terms of better understanding the prevalence of integrase inhibitor (INSTI) drug resistant mutations among ART patients who are virally unsuppressed.

### Limitations

Neither ART adherence nor the length of time an individual has been on ARVs were considered in this analysis. ART adherence was self-reported by participants, but these data were not used in this analysis due to possible social desirability bias. ART use was determined by ARVs detected in the blood specimen collected at the time of the survey, and detection is affected by the half-life of the medication of interest, where adherence is suboptimal. The number of participants included in the *ARV undetected* cohort who may have been unaware of their HIV status was not taken into account for this analysis. Some variables had to be discarded in the multivariable models due to the large reduction in the dataset’s size due to missingness, despite the univariate analysis finding of significant associations with the outcome variable.

## Conclusions

Trends of gender disparities in HIV program participation in the southern African region require further study. Analyzing comparable survey data across the region allows for an evaluation of possible obstacles that discourage men from participating in HIV programming and retention of care. Further insights are needed as to which factors are at play that encourage or act as barriers to treatment for men, but not for women.

Similar to antenatal and prevention of mother-to-child transmission (PMTCT) programs which have helped in initiating women on ART, there should be greater social investment in initiatives that solely target men. Identifying and adapting such interventions that can be effective at linking men to ART and improve knowledge about HIV treatment, will help reduce the national burden of disease and enable South Africa, a country with disproportional burden of infection, to finally reach epidemiological control.

## Data Availability

The datasets analyzed during the current study are available in the Human Sciences Research Council (HSRC) data research repository, http://datacuration.hsrc.ac.za/.

## Acknowledgement

The authors would like give thanks and acknowledgement to the entire SABSSM V field staff team, the field supervisors, the nurses, and the data collectors who worked tirelessly to conduct this survey.

## Disclaimer

The findings and conclusions in this manuscript are those of the authors and do not necessarily represent the official position of the funding agencies.

